# Spatial Distribution and Associated Factors Influencing 2024 Measles-Rubella Vaccination Campaign Coverage among Children Aged 9–59 Months in Mainland Tanzania

**DOI:** 10.1101/2025.08.01.25332784

**Authors:** Hajirani M. Msuya, Samwel Lwambura, Ibrahim Msuya, Omar Lweno, Bakari Fakih, Mwifadhi Mrisho, Hassan Tearish, Gumi Abdallah, Selemani Mmbaga, August Kuwawenaruwa, Moza Kassim, Kamaria Kassim, Farida Hassan, Joseph Mdachi, Furaha Kyesi, William Mwengee, Abdallah Mkopi

## Abstract

**Introduction:** Globally, measles remains a major cause of child mortality, and rubella is the leading cause of birth defects among all infectious diseases. In Mainland Tanzania, eliminating measles and rubella remains challenging due to geographical diversity, uneven healthcare facilities distribution, and socio-economic disparities across regions. This study aimed to explore the spatial distribution and associated factors of measles-rubella campaign coverage among children aged 9–59 months in Mainland Tanzania.

**Methods:** A cross-sectional survey was conducted to assess the spatial distribution and factors influencing measles-rubella (MR) vaccination coverage among children aged 9–59 months in mainland Tanzania after the MR campaign implementation in February 2024. The spatial autocorrelation was used to assess vaccination coverage in a study area. Hotspot and cold-spot analysis were performed to describe the spatial cluster of measles-rubella vaccination campaign coverage in Mainland Tanzania based on Getis-Ord Gi* statistics. To identify factors associated with the campaign coverage, we used a multivariable logistic regression model.

**Results:** The study involved 16,703 children, of whom 81.5% received the MR vaccine during the campaign. Vaccination coverage varied notably between regions, with Tabora and Pwani having a low coverage rate of 58.8% (95% CI: 51.3%-65.9%) and 61.0% (95% CI: 45.0%-75.0%) respectively. The Njombe and Mbeya demonstrating a high coverage rate of 97.4% (95% CI: 90.5%-99.3%) and 95.6% (95% CI: 90.9%-97.9%) respectively. The household wealth quintile and place of residence, caregiver’s education, caregiver’s age, and their marital status were associated with receiving MR vaccination during the campaign among children aged 9-59 months in Mainland Tanzania. Spatial distribution revealed significant clustering of vaccination coverage (Moran’s I = 0.34, p < 0.01). Hotspot analysis identified eight clusters with low coverage in —Tabora (five clusters) and Pwani (three clusters)— indicating higher proportions of unvaccinated children. Regions like Njombe and Mbeya were hotspots identified clusters with high vaccination coverage.

**Conclusion:** This study highlights critical gaps in measles-rubella vaccination coverage in Mainland Tanzania, where regional disparities hinder the achievement of global targets. It stresses the importance of the caregiver’s education and household wealth, advocating for targeted interventions and localized research to address these challenges. To drive progress, integrating geospatial artificial intelligence (GeoAI), spatial data science, and satellite technology is crucial. These advanced tools enable real-time, high-resolution mapping, optimizing resource allocation, and identifying underserved areas. By leveraging these technologies, we can ensure data-driven, efficient interventions that leave no region behind, ultimately achieving universal vaccination coverage and protecting every child globally.

## Background

Measles, a highly contagious viral disease, remains a leading cause of vaccine-preventable deaths among young children globally, despite the availability of an effective vaccine [1]. The measles vaccine has significantly reduced cases and deaths worldwide, but coverage gaps remain, particularly in sub-Saharan Africa, where infrastructure, healthcare access, and socio-economic factors hinder vaccination efforts [2, 3]. Additionally, rubella infection during pregnancy can lead to congenital rubella syndrome (CRS), resulting in significant disability and health costs [4]. Both diseases remain critical public health challenges, with measles and rubella elimination efforts often combined in a two-dose vaccination approach as recommended by the World Health Organization (WHO) [5].

In Tanzania, efforts to eliminate measles and rubella have led to the implementation of various vaccination campaigns targeting children under five years old [6, 7]. The Measles-Rubella (MR) vaccination campaign is essential for reducing transmission, but achieving optimal coverage remains challenging due to Tanzania’s geographical diversity, uneven healthcare distribution, and disparities in socio-economic factors across regions [8]. Prior assessments of MR vaccination coverage in the year 2019 have shown substantial variations across the country, at the national level, coverage of MR was at 88.2%, with MR coverage varying across regions from 69.0% in Katavi to 100% in Kusini Unguja [9]. Despite high national coverage, some regions still have yet to reach optimal coverage with some councils receiving less than 80% [9]. The recently study conducted in Tanzania reported measles outbreak and was confirmed in July 2022, as per the WHO Measles Outbreak Guide (2022) [10]. The most affected age group were children less than 5 years and 7-15 years age groups. The outbreak was first confirmed in Dar es Salaam and then spread to involve Tanga and Pwani, which are all along the coast. In Zanzibar, the measles outbreak was also confirmed in two districts in Unguja and three districts in Pemba with 20 laboratory-confirmed cases [11].

To maintain the effectiveness of the previous MR campaign 2019 and responding to recently measles outbreak, the Ministry of Health (MoH) in Tanzania, conducted a follow-up MR campaign in February 2024. The 2024 measles-rubella survey aims to improve vaccine coverage among children aged 9-59 months, targeting a critical age group for both primary measles and rubella vaccination. Using 2024 measles-rubella survey data to identify clusters of low vaccination coverage as well as understanding factors influencing this coverage are crucial for optimizing the campaign’s effectiveness especially in underserved and high-risk areas [2]. Spatial distribution studies highlight that geographical barriers, socio-economic factors, marital status, higher poverty levels, parental education, and rural-urban divides, significantly impact of measles-rubella vaccine accessibility [12–14]. Addressing these barriers requires a multifaceted approach, including targeted health education and improved healthcare delivery systems [15], particularly in high-risk areas identified through spatial distribution [16].

The spatial distribution with regards to measles-rubella campaign coverage survey, the researchers begin by assessing the unique characteristics of different areas, including population density, vaccination rates, and healthcare access, as these factors significantly impact vaccination coverage. After identifying these factors, spatial relationships are analyzed to identify coverage patterns across regions [17]. Common techniques include disease mapping, spatial clustering, and risk factor identification through map comparisons [18]. Spatial clustering is particularly effective in pinpointing areas with low vaccination coverage, which are often at higher risk for outbreaks due to shared environmental or socio-economic factors influencing vaccine access and uptake [19]. Cuzick and Edwards (1990) identified three methods for detecting spatial clustering: cell count data, autocorrelation of neighboring cells, and analyzing the distance between vaccination events or coverage rates [20]. Advanced numerical methods like scan statistics [21], statistics by Ohno et al [22], Poisson statistics [23], Global Moran’s I [24], Global Geary’s C [25], General Getis-Ord’s G [26], and Local Moran’s I [27] are critical for detecting spatial clusters in health surveys. Hotspot analysis using Getis-Ord Gi* statistics (pronounced G-i-star) reveals clustering intensity, with positive scores indicating hotspots —areas with high coverage and negative scores showing cold spots —areas with low coverage [28].

Several studies have shown that mapping spatial distribution can effectively guide vaccination strategies, allowing public health officials to prioritize interventions in underserved areas and optimize resource allocation [18, 29]. In Tanzania, as in other countries, utilizing spatial data to assess measles-rubella vaccine campaign coverage can play a crucial role in: i) identifying coverage gaps, ii) uncovering factors that drive disparities in vaccine coverage across various areas, iii) tackling spatial and demographic inequities in vaccine distribution, and iv) enabling more focused and efficient vaccination efforts [2, 29, 30]. Therefore, this study aimed to investigate the spatial distribution and factors influencing 2024 measles-rubella campaign coverage among children aged 9–59 months in mainland Tanzania.

## Methods

### Study Design and Setting

A cross-sectional survey was conducted employed the quantitative methods used to assess coverage achieved during the 2024 measles-rubella campaign in mainland Tanzania. The quantitative data were collected by interviewing caregivers of children 9-59 months within the selected households. The household was defined as a group of individuals who share the same cooking pot or kitchen for food preparation. The main target population for the post-campaign evaluation was children aged 9-59 months. The survey was done in all regions of the mainland Tanzania after the campaign implementation in February 2024.

### Sample Size

The sample size was determined to ensure accurate, age- and sex-specific MR vaccine coverage estimates for children aged 9–59 months at the national level. With a target precision of ±5% and an expected coverage rate of 95%, a total of 14,040 households were required for the sample size.

### Sampling Procedures

The study employed a two-stage stratified cluster sampling approach [31]. The first stage involved selecting Primary Sampling Units (PSUs) or enumeration areas (EAs) from Tanzania National Bureau of Statistics (NBS). PSUs were selected randomly using a probability proportional to size (PPS) method to ensure that larger population areas had a higher chance of selection, leading to more accurate and representative results. These PSUs represented the various regions of Mainland Tanzania, capturing a diverse demographic for the study. In the second stage, Secondary Sampling Units (SSUs) were selected within each EA. SSUs were households with at least one child aged 9–59 months, the target population for the measles-rubella campaign. In this study, each EA was listed, and a sketch map of the area was created to accurately locate the selected households. Households were assigned identification numbers for tracking.

To randomly select households, a randomizer tool was used to choose eight households per EA. If a household could not be located or did not meet the eligibility criteria, no replacement was made, and those households were excluded from the study. Additionally, households that did not provide consent or could not be accessed after three extended visits were excluded from the final sample. All eligible caregivers aged 18 or older in the selected households were interviewed. This included both regular residents and visitors who slept in the household the night before the survey. Interviewers used the household identification number and sketch map to locate and confirm the eligible households. The interviews focused on gathering data regarding vaccination status, demographic factors, and any barriers to vaccine access to understand the spatial patterns of vaccine coverage, factors contributing to coverage disparities, and the effectiveness of the 2024 measles-rubella campaign.

### Data Collection

A total of 5 scientists from IHI, 26 regional supervisors, and 84 interviewers from all 26 regions in Mainland Tanzania attended training and a pre-test in Dar es Salaam. An average of 3 interviewers were selected for each region from the IHI database. The interviewers and regional supervisors were trained to understand the survey objectives, importance, and questionnaire. They were also trained in interview techniques to maximize response rates and ensure data validity. The roles of each study team member were clarified during the training. A training manual was prepared to enhance training effectiveness and serve as a reference during data collection. The study instruments were pilot-tested in an excluded EA to ensure understanding, correct wording of questions, and capture the intended data. Pilot testing also helped to estimate survey duration and confirm the survey teams’ understanding of sampling households in the field.

All quantitative data were collected by well-trained interviewers who have experience in administering quantitative survey studies. The data were collected using mobile devices (tablets) that were programmed with the Open Data Kit (ODK) application. All eligible respondents were interviewed in their households. In each sampled household, the primary caregiver of children aged 9 to 59 months. For the evaluation of the MR vaccine, structured questionnaires were used as tools to collect data. These questionnaires assessed the vaccination status for MR, key demographic information, household socio-economic status (SES), demand-side constraints, and perceptions about the campaign. Interviews were conducted with caregivers of eligible children to determine if their children had received the MR vaccine during the campaign.

The data collectors were required to check if the left-hand little fingers of the children were marked with special ink during the campaign as a certification of vaccination. If there was no mark, caregivers were asked to recall whether their children had received the MR vaccine by inquiring about the specific MR vaccination procedure. The MR vaccination procedure followed these steps: caregivers were asked if they remembered whether their children received the measles-rubella vaccine during the campaign. If yes, they were prompted to recall when and where the vaccination took place. Caregivers were also asked if they were informed or reminded about the vaccine during the campaign. Additionally, caregivers were inquired whether their child received the vaccine via injection in the left shoulder during the campaign period. Clinic card marks were also used as part of the assessment process.

### Data Quality Control

Quality control was vital for this assignment, with the Ministry of Health (MoH) staff collaborating closely with the senior study team during fieldwork. A skilled research team conducted the survey, using detailed field tools developed in partnership with MoH staff, which were piloted in training sessions. Each team had a field supervisor ensuring daily quality control, with supervisors accompanying at least one interview daily and randomly reviewing later interviews. Completed questionnaires were checked daily by interviewers and supervisors. The research team provided close oversight during training, data collection, entry, cleaning, and validation. Initial data cleaning was performed by interviewers on tablets before online uploads, with data clerks correcting errors using a user guide. A second data cleaning level occurred post-download by the Data manager, involving interviewers for clarification on discrepancies. Daily field reports from the regional data manager/supervisor further supported the quality control process.

### Primary outcome and explanatory variables

In this study the outcome variables were measles-rubella campaign coverage for children aged 9–59 months coded as “Yes” if their children had received the MR vaccine during the campaign and “No” if the child failed to receive the vaccine. Information on vaccination status during the campaign was obtained in the following ways; initially, finger markers were intended to be used to verify the coverage. The Post-MR campaign vaccination coverage survey was conducted in June, 2024 throughout mainland Tanzania, after the campaign implementation in February 2024, but verification of children’s vaccinations using finger markers was not possible due to significant delays in fieldwork. As a result, historical information or the caregiver’s recall was relied upon to assess coverage, supplemented by follow-up questions to confirm the caregivers’ awareness of the marked finger and the method of vaccine administration. The predictor’s variables were the caregiver’s age, marital status, sex of household head, sex of the child, caregiver’s education level, household size, occupation, wealth index, residence, and regions.

### Data Management and Data Analysis

Data extraction, cleaning, weighting, and recording were done before any statistical analysis. Descriptive statistics such as cross-tabulations and frequency tables were used to describe the characteristics of the study population.

### Spatial and Hotspots Analysis

In this study, spatial autocorrelation was applied to assess vaccination coverage in the area. The Moran’s Index (Moran’s I) measured spatial patterns, determining if coverage was clustered, dispersed, or random [32], with significant values (p < 0.05) confirming spatial patterns [33]. Hotspot analysis using the Getis-Ord Gi* statistic was applied in this study to identified areas of high and low measles-rubella vaccination coverage. The Getis-Ord Gi* statistics use the z-scores, the positive z-scores indicated the hotspots —areas with high MR campaign coverage, while the negative z-scores highlighted the cold spots— areas with low MR campaign coverage. The Getis-Ord Gi* statistics values for hot and cold spots are approximately *±*1.96 and *±*2.58 at the 5% and 1% significance levels, respectively [26–28]. The hotspot measles-rubella vaccination coverage in this study was visualized with blue (cold spots) and green (hotspots).

### Logistic Regression Analysis

To identify factors associated with the campaign coverage, we used a multivariable logistic regression model. After fitting a bi-variable logistic regression analysis, variables with a *p-value* of *<* 0.2 in the bi-variable analysis were further considered in the multivariable logistic regression analysis. *Adjusted odds ratio* (AOR) with 95% *CI* and *p-value <*0.05 in the multivariable logistic regression model were used to declare statistically significant variables associated with campaign coverage. In this study, we use the R software [34] to execute the data analysis.

### Ethics statement

This study was conducted by an experienced research team, ensuring that national, regional, and district health authorities were provided with sufficient information about the study. Written informed consent was obtained from the head of each household and translated into Swahili (Tanzania national language) to explain the purpose, the researchers involved, and the study’s details. The translated consent was also used to introduce the survey team to community leaders. The study followed the International Ethical Guidelines for Biomedical Research Involving Human Subjects [35], as recommended by the WHO. Verbal informed consent was used for a standard coverage survey like this, as it was non-intrusive and did not involve sensitive information. A standard explanation and introduction were provided to each caregiver before seeking individual informed consent through a proxy. Caregivers were allowed to ask questions and indicate their willingness to participate in the survey. The confidentiality of all participants was ensured, and the surveys posed no risk to them. Any participant with an acute illness was referred to a healthcare facility. Ethical approval was obtained from the Ethical Committee of Ifakara Health Institute (IHI/IRB/No: 14-2024).

## Results

A total of 16703 caregivers with children aged 6-59 months participated in this study. Thirteen thousand six hundred and two (81.5%) of the children received measles-rubella vaccination during the campaign. There is an equal proportion in terms of sex of children who received measles-rubella vaccination during the campaign. The majority of children’s received the measles-rubella vaccine lived with caregiver’s who attained primary education level. Eight thousand nine hundred and thirty-five (65.7%) of the vaccinated children during the campaign lived in rural areas. The majority of the surveyed caregiver’s were married/cohabited. In term of wealth, 5621 (41.3%) of the vaccinated children during the campaign came from the middle (Table 1)

**Table 1:** Characteristics of study participants and MR coverage by various background characteristics among children aged 9–59 months in Tanzania.

| Variables | Received MR |  |  | p-value<sup>2</sup> |
| --- | --- | --- | --- | --- |
|  |  | No | Yes |  |
|  | Overall, N = 16703<sup>1</sup> | n = 3101<sup>1</sup> | n = 13602<sup>1</sup> |  |
|  | N (%) | n (%) | n (%) |  |
| Child sex |  |  |  | 0.12 |
| Male | 8344 (50.0) | 1588 (51.2) | 6756 (49.7) |  |
| Female | 8359 (50.0) | 1513 (48.8) | 6846 (50.3) |  |
| Caregiver's age |  |  |  | <0.001 |
| 18 – 24 | 2819 (16.9) | 603 (19.4) | 2216 (16.3) |  |
|  |  | No | Yes |  |
|  | Overall, N = 16703<sup>1</sup> | n = 3101<sup>1</sup> | n = 13602<sup>1</sup> |  |
|  | N (%) | n (%) | n (%) |  |
| 25 – 34 | 7864 (47.1) | 1488 (48.0) | 6376 (46.9) |  |
| 34+ | 6020 (36.0) | 1010 (32.6) | 5010 (36.8) |  |
| Caregiver’s education |  |  |  |  |
| No formal education | 2256 (13.5) | 563 (18.2) | 1693 (12.4) |  |
| Primary education | 10942 (65.5) | 1937 (62.5) | 9005 (66.2) |  |
| Secondary education and more | 3505 (21.0) | 601 (19.3) | 2904 (21.4) |  |
| Residence |  |  |  | <0.001 |
| Rural | 10858 (65.0) | 1923 (62.0) | 8935 (65.7) |  |
| Urban | 5845 (35.0) | 1178 (38.0) | 4667 (34.3) |  |
| Caregiver’s marital status |  |  |  | 0.14 |
| Single/Not in union | 788 (4.7) | 137 (4.4) | 651 (4.8) |  |
| Married/Cohabited | 14093 (84.4) | 2603 (83.9) | 11490 (84.5) |  |
| Separated/Divorced | 1395 (8.4) | 288 (9.3) | 1107 (8.1) |  |
| Widowed | 427 (2.6) | 73 (2.4) | 354 (2.6) |  |
| Caregivers sex |  |  |  | 0.6 |
| Male | 588 (3.5) | 104 (3.4) | 484 (3.6) |  |
| Female | 16115 (96.5) | 2997 (96.6) | 13118 (96.4) |  |
| Wealth status |  |  |  | <0.001 |
| Poor | 3600 (21.6) | 840 (27.1) | 2760 (20.3) |  |
| Middle | 6774 (40.5) | 1153 (37.2) | 5621 (41.3) |  |
| Rich | 6329 (37.9) | 1108 (35.7) | 5221 (38.4) |  |

<sup>1</sup>n (%), <sup>2</sup>Pearson's Chi-squared test

### Measles-Rubella Vaccination Campaign Coverage in Mainland Tanzania

In mainland Tanzania, an estimated 81.5% (95% CI: 80.0%-82.9%) of the children aged 9–59 months received the MR campaign dose, as assessed by caregivers’ recall. The coverage varied among different regions, with Tabora and Pwani having a low coverage rate of 58.8% (95% CI: 51.3%-65.9%) and 61.0% (95% CI: 45.0%-75.0%) respectively. The Njombe and Mbeya demonstrated a high coverage rate of 97.4% (95% CI: 90.5%-99.3%) and 95.6% (95% CI: 90.9%-97.9%) respectively. The estimates of the measles-rubella campaign coverage are presented in Table 2 and Figure 1.

**Figure 1.**
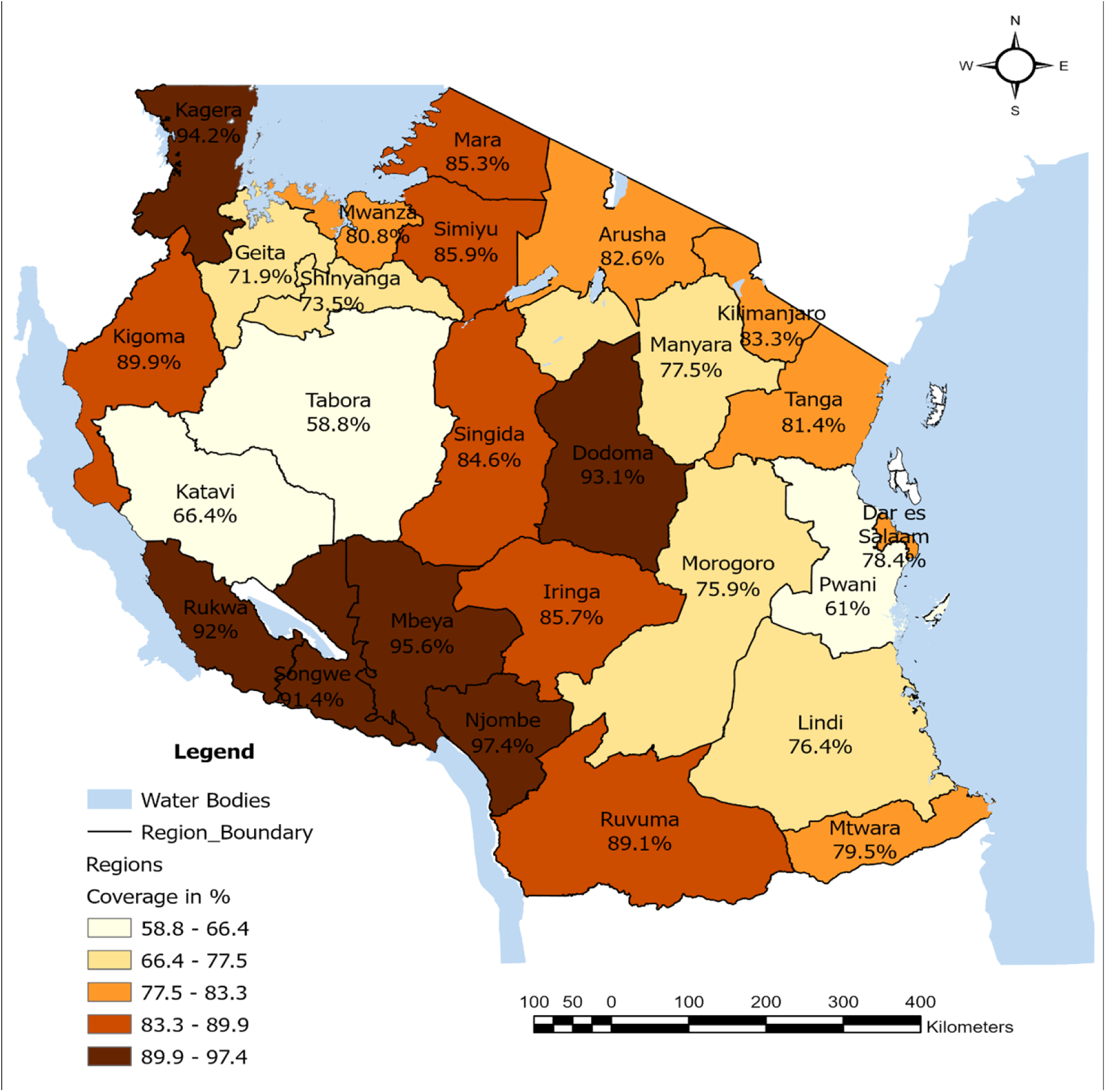
Map of MR vaccination rate among children aged 9–59 months across regions of Tanzania, MR survey 2024.

**Table 2:** Estimated measles-rubella campaign vaccination coverage – caregiver’s recall.

| Region | Weighted % | 95%CI | n | N |
| --- | --- | --- | --- | --- |
| Njombe | 97.4 | 90.5-99.3 | 167 | 172 |
| Mbeya | 95.6 | 90.9-97.9 | 570 | 601 |
| Kagera | 94.2 | 90.5-96.5 | 1,005 | 1072 |
| Dodoma | 93.1 | 90.5-95.0 | 709 | 777 |
| Rukwa | 92.0 | 85.6-95.6 | 329 | 363 |
| Songwe | 91.4 | 80.4-96.5 | 286 | 307 |
| Kigoma | 89.9 | 83.6-94.0 | 763 | 864 |
| Ruvuma | 89.1 | 82.8-93.3 | 267 | 298 |
| Simiyu | 85.9 | 80.4-90.0 | 1,032 | 1213 |
| Iringa | 85.7 | 76.4-91.7 | 143 | 168 |
| Mara | 85.3 | 80.4-89.2 | 893 | 1033 |
| Singida | 84.6 | 76.5-90.3 | 375 | 460 |
| Kilimanjaro | 83.3 | 75.7-88.9 | 401 | 478 |
| Arusha | 82.6 | 75.7-87.9 | 394 | 462 |
| **Overall** | **81.5** | **80.0-82.9** | **13,602** | **16703** |
| Tanga | 81.4 | 74.2-87.0 | 422 | 513 |
| Mwanza | 80.8 | 75.2-85.5 | 1,093 | 1346 |
| Mtwara | 79.5 | 72.5-85.1 | 298 | 365 |
| Dar Es Salaam | 78.4 | 74.3-82.1 | 889 | 1163 |
| Manyara | 77.5 | 68.9-84.3 | 435 | 564 |
| Lindi | 76.4 | 52.5-90.5 | 196 | 232 |
| Morogoro | 75.9 | 67.2-82.9 | 507 | 647 |
| Shinyanga | 73.5 | 63.8-81.3 | 444 | 643 |
| Geita | 71.9 | 65.2-77.8 | 814 | 1168 |
| Katavi | 66.4 | 48.8-80.4 | 249 | 343 |
| Pwani | 61.0 | 45.0-75.0 | 188 | 272 |
| Tabora | 58.8 | 51.3-65.9 | 733 | 1179 |

#### Spatial Autocorrelation *and* Hotspot analysis of MR campaign coverage

The spatial distribution of MR coverage in Mainland Tanzania was found to be non-random (*Moran’s I* = 0.34, *p-value* < 0.01). The Moran’s Index value observed (0.34) exceeded the expected value (−0.004), and the p-value was statistically significant (<0.01). The results from the *Getis-Ord GI\** statistical analysis revealed most of the cold spots (areas with low MR campaign coverage) were found in the western zone containing Tabora, and Katavi. Some of the cold spots were also portrayed in the Lake Zone, Shinyanga as well as the eastern zone parts including Pwani and Dar-es Salaam. In Tabora, areas with notably low coverage included Kigwa, Mwantundu, Kitangili, Bukoko, and Tambukareli. Similarly, in Pwani, the most notable low-coverage hotspots were in Msata, Makurunge, and Chole.

The map shows hotspots (areas with high MR campaign coverage) in the southern zone; region including Mbeya and Njombe and in the small pocket of the Lake zone, Kagera part of Tanzania. Conversely, Njombe and Mbeya were recognized as significant hotspots, indicating areas with higher MR campaign coverage. This suggests that children in these regions were more likely to receive the MR vaccine during the campaign. The description of the hotspots and cold spot are presented in Figure 2.

**Figure 2.**
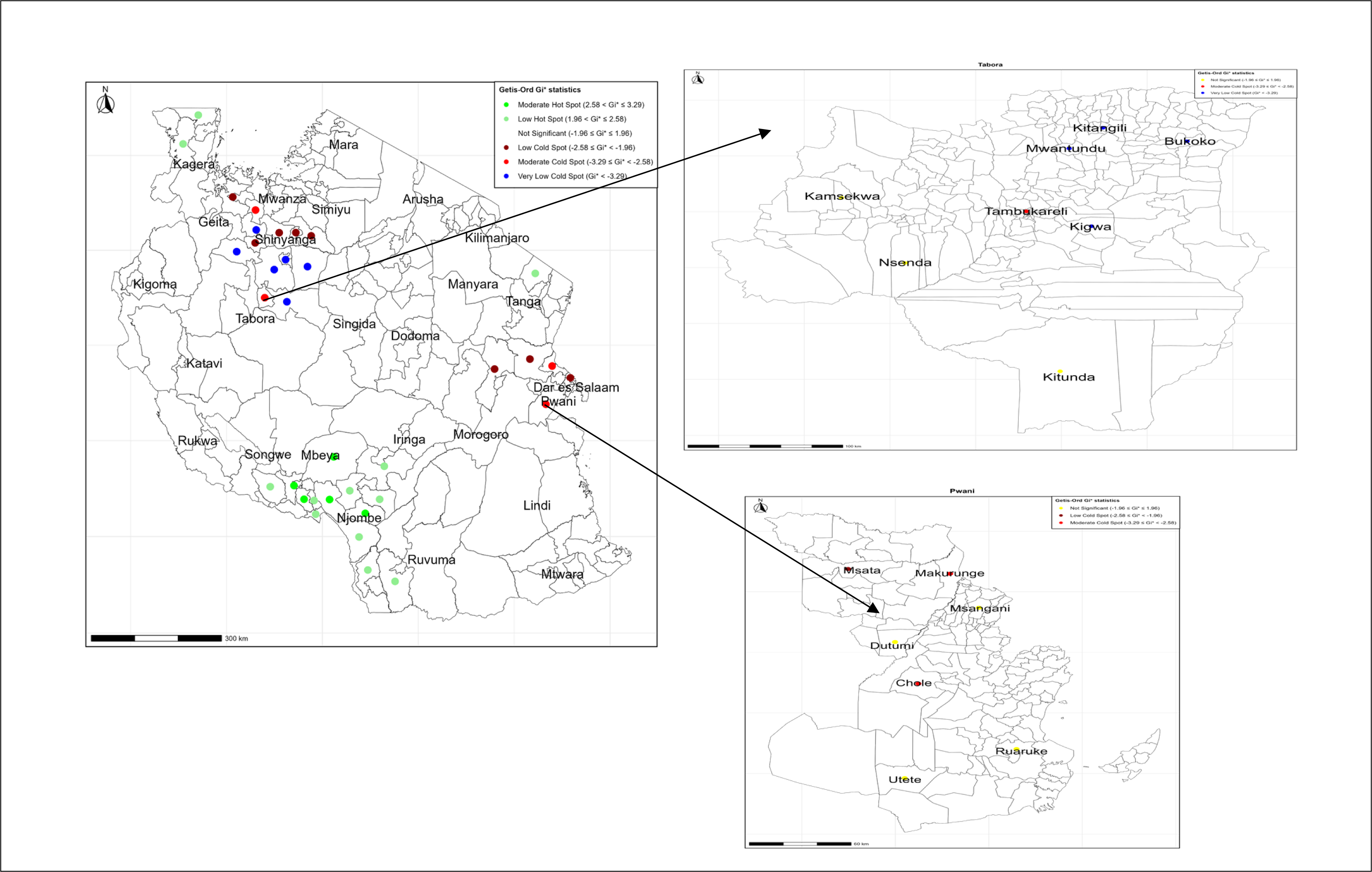
The spatial distribution of low measles-rubella (MR) vaccination coverage hot and cold spots in Tanzania Mainland, based on Getis-Ord Gi* statistical analysis of MR Survey 2024. The left panel shows regional-level clustering across the country, while the right panels display sub-regional clustering within Tabora (top) and Pwani (bottom) regions.

#### Factors associated with MR campaign coverage

In the bi-variable analysis, variables such as the caregiver’s age, caregiver’s education, household wealth status, region, and place of residence were associated with receiving the MR vaccine during the campaign (*p-value* < 0.2). Hence, these variables were eligible for the multivariable logistic regression. However, in the multivariable logistic regression analyses, only the caregiver’s education, caregiver’s age, household wealth status, region, and place of residence were significant at *p-value* < 0.05.

Among the individual level factors; the odds of receiving MR vaccination during the campaign for children who lived with caregivers who had primary school education were 1.4 (AOR = 1.40, 95% CI: 1.13-1.74) times higher compared to those children who lived with caregivers who did not attain any formal education. Children who lived with caretakers with secondary or higher education were approximately 1.4 (AOR = 1.44, 95% CI: 1.12 - 1.85) times more likely to have MR vaccine during the campaign than children who lived with caretakers with no formal education. The odds of receiving the MR vaccination during the campaign for children lived with caregivers aged 25 to 34 years were 1.2 (AOR = 1.20, 95% CI: 1.00 - 1.45) times more likely than those of children who lived with caregivers aged 18 to 24 years. Children who lived with caregivers aged 35 years and above were 1.6 (AOR = 1.55, 95% CI: 1.27 - 1.89) times more likely to have MR vaccine during the campaign than children who lived with caregivers aged 18 to 24 years. Children born to the wealth households were approximately 2 (AOR = 1.82, 95% CI: 1.34 - 2.43) times more likely to have MR vaccine than children born to poorest households.

Among community-level factors, children in the Tabora region had a 69% (AOR = 0.31, 95% CI: 0.19 - 0.51) lower chance of receiving the MR vaccination during the campaign than children in Arusha region. In contrast, children in the Njombe region had 7 (AOR = 6.77, 95% CI: 1.76 - 26.0) times high likelihood of receiving the MR vaccination during the campaign than children in Arusha region. The odds of receiving MR vaccination during the campaign among children in the urban resident were decreased by 37% (AOR = 0.63, 95% CI: 0.51-0.77) compared to those children who were rural areas. (Table 3).

**Table 3.**
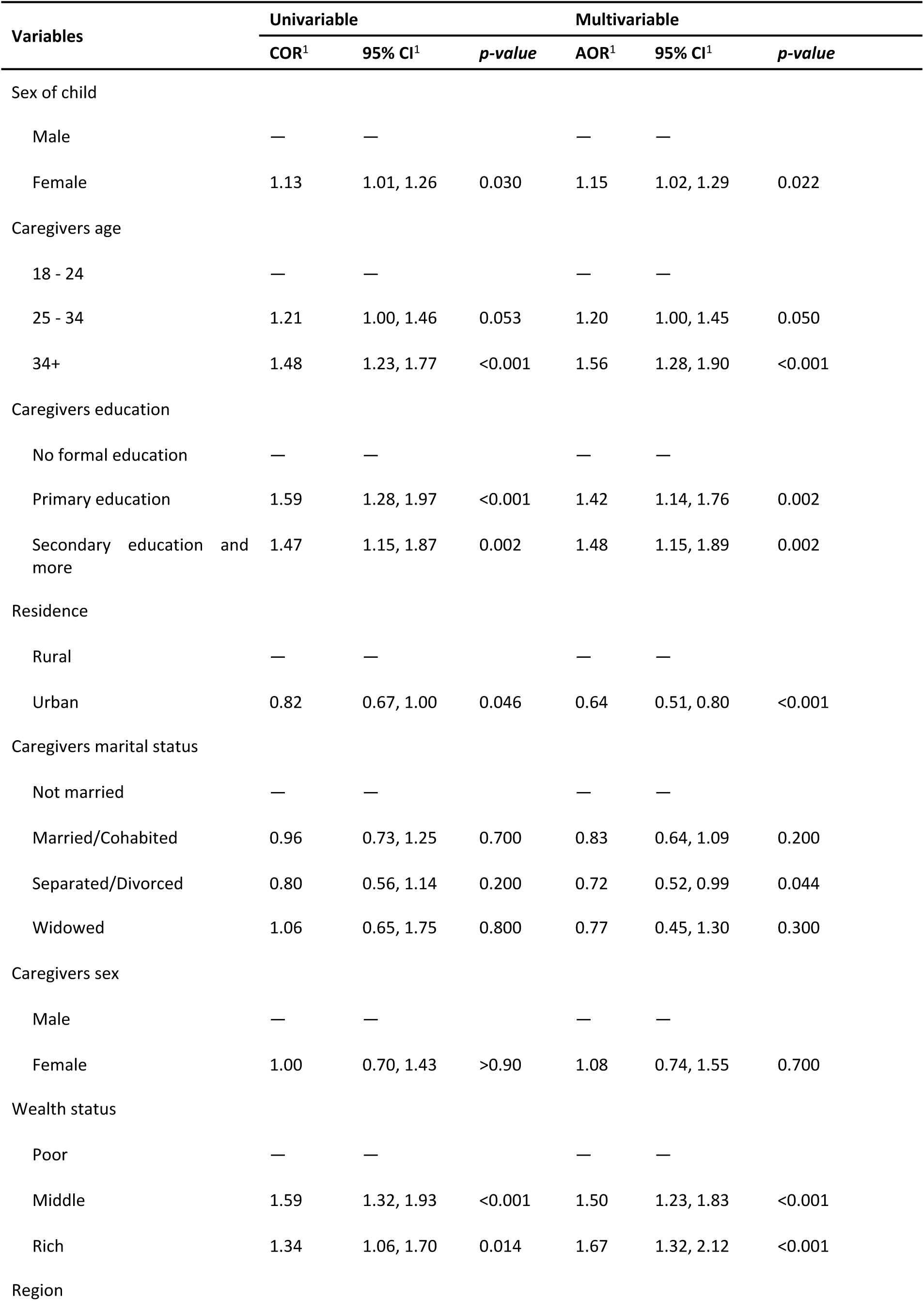
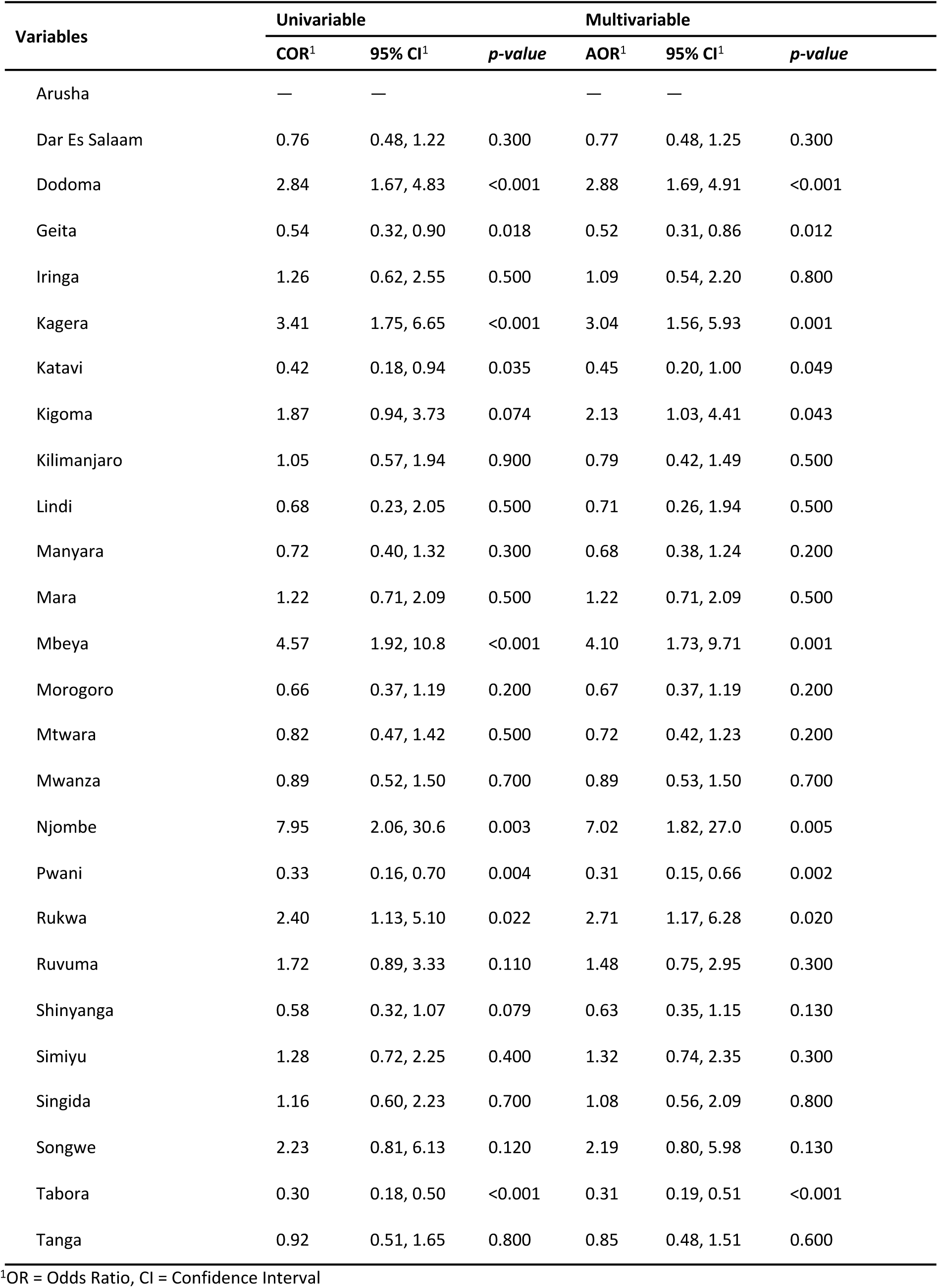
Multivariable multilevel logistic regression analysis of individual and community-level factors associated with measles vaccination among children aged 9– 59 months in Tanzania.

## Discussion

This study revealed that overall 81.5% of children aged 9–59 months in Mainland Tanzania had received the measles-rubella vaccine during the campaign which is lower than the previous coverage of the MR vaccination in 2019 which was 88.2% [9]. The coverage of the 2024 MR vaccine did not meet the global target of 95% or higher [36] of the population must be vaccinated [37, 38], means many targeted children were not reached during the MR vaccination campaign in Mainland Tanzania. However, we demonstrated that the overall measles-rubella coverage among children aged 9–59 months in Mainland Tanzania was low as global target [36], which is similar to other low- and middle-income countries (LMICs) such as Ethiopia (58.8%), but lower than Afghanistan (66%) and Pakistan (76%) [39]. A possible explanation on this disparity in vaccination coverage is likely influenced by a range of factors, both distinct and cross-cutting. Notably, barriers such as limited healthcare access in remote areas, vaccine supply interruptions, insufficient public awareness, cultural opposition, and inadequate outreach to marginalized communities contribute significantly. Cross-cutting issues, including widespread vaccine mistrust driven by misinformation, economic constraints like transportation costs, and ineffective communication strategies, further exacerbate vaccine hesitancy.

In this study, our findings show that the measles-rubella vaccination coverage was highest in Njombe region which was 97.4%, and lowest in the Tabora region which was 58.8%. This result is supported by previous studies in Tanzania that showed childhood vaccination coverage varied across different regions of the country [9, 40]. The 2022 Tanzania Demographic and Health Survey and Malaria Indicator Survey reported a national decline in full vaccination against all basic antigens coverage for children aged 12–23 months, from 75% in the year 2015–16 to 53%, with regional coverage ranging from 3% in Shinyanga to 66% in Kilimanjaro [41]. The disparity between Njombe and Tabora underscores the need for focused interventions and resource allocation. Improving vaccination coverage in underperforming regions can reduce measles and rubella burdens, contributing to better public health and advancing global vaccination goals.

Donors and policymakers are increasingly interested in mapping the spatial heterogeneity of childhood vaccination [42] to identify existing gaps and intervene accordingly. The spatial autocorrelation analysis of our data showed a clustering pattern of childhood measles vaccination coverage were mostly western, eastern, lake zone part of Tanzania. Studies in various countries, including Ethiopia [43], Mozambique [44], and Brazil [45], reveal significant spatial heterogeneity in vaccination coverage. For example, Ethiopia’s research showed clustering in regions like Afar and Amhara, while Mozambique identified regional disparities with lower coverage. In São Paulo, Brazil, varied measles vaccination rates were observed across municipalities. Contrastingly, the Somali Region in Ethiopia had lower coverage in zones like Nogob and Erer [46]. Factors like healthcare infrastructure, cultural practices, and data collection could explain these discrepancies. Tanzania needs further research to understand these clustering patterns, and collaboration with neighboring countries could inform targeted interventions. In addition; to address these disparities, targeted interventions must be tailored to the specific geographical, economic, and social conditions of each region.

In the multilevel analysis, caregiver’s age, caregiver’s education, household wealth status, region, and place of residence were significantly associated with measles-rubella immunization. Children lived with caregiver’s who had primary school education and secondary and above school education had higher odds of receiving measles-rubella vaccine during the campaign than children lived with caregiver’s or uneducated mothers. This is supported by previous studies done in Ethiopia [47], the Democratic Republic of Congo [48], and China [49]. A possible explanation might be that caregiver’s maternal education is vital to enhance awareness about childhood vaccination and to improve the utilization of primary health-care services, such as childhood vaccination services. Educated caregiver’s tend to have improved communication skills, which makes interactions with health-care providers easier, and leads to a better comprehension of vaccination programs and practices.

Across the world, one common theme emerges in the fight against measles and rubella: socio-economic status plays a crucial role in vaccination coverage. In Vietnam [50], children from lower-income families, living in rural areas, and with less-educated mothers, were less likely to receive timely immunizations. Similarly, in Japan [51], higher income inequality led to lower vaccination rates, while communities with stronger social ties saw better coverage. Tanzania [9], Niger [52], Canada [53], and India [54] all echoed this pattern, with wealthier families more likely to vaccinate their children. The World Health Organization (2023) confirms that inequities in immunization, driven by socio-economic factors, continue to hinder global efforts to eliminate these preventable diseases [55]. The story is clear: addressing these disparities is key to ensuring every child, regardless of background, is protected from measles and rubella.

Place of residence significantly influences measles-rubella vaccination coverage, with urban areas generally exhibiting higher vaccination rates than rural counterparts. This disparity is attributed to factors such as improved healthcare infrastructure, better access to vaccination services, and increased health awareness in urban settings. Our results revealed that, children in urban setting had lower chance of receiving the MR vaccination during the campaign compared to those children who lived in rural areas. This is contradicting to a study in Ethiopia found that urban children had a 28% higher full vaccination coverage compared to rural children [56]. Conversely, research in Pakistan which revealed that children residing in remote rural areas had higher measles vaccination coverage compared to urban areas, with rates of 89.8% in rural areas versus 75.2% in urban areas [57]. These contrasting findings underscore the complex relationship between place of residence and vaccination coverage, suggesting that factors such as healthcare accessibility, cultural practices, and local health policies play significant roles in vaccination rates.

This study has its strengths and limitations. This study was strong in a number of ways. The research team collected data from a variety of sources, such as vaccination cards, medical records, and mother/caregiver recall, were used to determine childhood vaccination coverage. The collection of high-quality data was also facilitated by the utilization of digital tools and the employment of skilled data collectors. However, the study also had limitations, including vulnerability to biases that are closely related to the cross-sectional study design, such as recall bias and non-response bias. Mothers or caregivers who lacked vaccination cards may have forgotten their child’s vaccination status, which could have led to misclassification. The survey was conducted 4-5 months after the MR vaccination campaign. The ink previously used to mark the fingers had already been removed, so the research team relied on the caregivers’ memory of events before and during the vaccine administration. However, this reliance on memory may have introduced some recall bias. To reduce this bias, research assistants reviewed asked the series of the questions related to the MR vaccine provision.

## Conclusion

This study reveals the pressing challenge of measles-rubella vaccination coverage in Mainland Tanzania, where significant regional disparities have resulted in coverage falling short of global targets. The findings underscore the pivotal role of caregiver education and household wealth in shaping vaccination outcomes, highlighting an urgent need for targeted interventions in regions facing the greatest challenges. Moving forward, localized research, addressing socio-economic barriers, and fostering meaningful community engagement will be essential to achieving global vaccination goals. Additionally, embracing the transformative potential of geospatial artificial intelligence (GeoAI), spatial data science, and satellite technology will be crucial in reshaping the landscape of vaccination efforts. By leveraging these cutting-edge technologies, we can optimize the allocation of resources and tailor public health interventions to local needs, ultimately reducing regional disparities by ensuring that no region is left behind in the fight to meet the global vaccination targets. Hence, bridge gaps in healthcare access and ensure a healthier, more equitable future for all.

## Data Availability

All relevant data are within the manuscript and its Supporting Information files.

